# Kidney Replacement Therapy in COVID-19-Related Acute Kidney Injury: the Importance of Timing

**DOI:** 10.1101/2024.01.13.24301272

**Authors:** Carlos Augusto Pereira de Almeida, Marcia Fernanda Arantes de Oliveira, Alexandre Macedo Teixeira, Carla Paulina Sandoval Cabrera, Igor Smolentzov, Bernardo Vergara Reichert, Paulo Ricardo Gessolo Lins, Camila Eleuterio Rodrigues, Victor Faria Seabra, Lucia Andrade

## Abstract

The objective of this study was to evaluate two different criteria for deciding when KRT should be initiated in patients with COVID-19-related AKI, as well as to determine the impact of the timing of KRT, as defined by each criterion, on in-hospital mortality among such patients. This was a retrospective study involving 512 adult patients admitted to the ICU. All participants had laboratory-confirmed COVID-19 and a confirmed diagnosis of AKI. The potential predictors were the determination of the timing of KRT based on a temporal criterion (days since hospital admission) and that based on a serum creatinine cutoff criterion. Covariates included age, sex, and the SOFA score, as well as the need for mechanical ventilation and vasopressors. The main outcome measure was in-hospital mortality. We evaluated 512 patients, of whom 69.1% were men. The median age was 64 years. Of the 512 patients, 76.6% required dialysis after admission. The overall in-hospital mortality rate was 72.5%. When the timing of KRT was determined by the temporal criterion, the risk of in-hospital mortality was significantly higher for delayed KRT than for timely KRT—84% higher in the univariate analysis (OR=1.84, 95%, [CI]: 1.10-3.09) and 140% higher after adjustment for age, sex, and SOFA score (OR=2.40, 95% CI: 1.36-4.24). When it was determined by the creatinine cutoff criterion, there was no such difference between high and low creatinine at KRT initiation. In patients with COVID-19-related AKI, earlier KRT appears to be associated with lower in-hospital mortality.

## Introduction

In Brazil, the first case of COVID-19 was reported in February 2020,^1,2^ and more than 36 million cases have been reported since then.^3^ The state of São Paulo alone has accounted for more than 6 million cases.^4^

Although COVID-19 mainly affects the respiratory system, some patients developing acute respiratory distress syndrome,^5-7^ we now know that it is not exclusively a respiratory disease.^8,9^ Early in the pandemic, some studies showed that kidney involvement was uncommon among patients with COVID-19.^10,11^ However, more than 40% of such patients develop proteinuria, hematuria, or both.^8,12^ Small studies from China, Europe, and the United States have reported a wide range of values for the incidence of acute kidney injury (AKI) in patients with COVID-19, from 1% and 42%.^13,14^ We now also know that AKI can occur in critically ill patients with COVID-19, as noted in 20-40% of those admitted to intensive care units (ICUs) in Europe and the United States.^15,16^

In the largest study to date of patients with COVID-19 in Latin America,^17^ AKI was found to be hospital-acquired in 64.7% of the patients and 46.2% of the patients required kidney replacement therapy (KRT). In Brazil, the mortality rate associated with COVID-19-related AKI was reported to be 72.5% in a multicenter study conducted in the city of São Paulo^18^ and 93.0% in a study conducted in the city of Recife.^27^ Nevertheless, COVID-19-related AKI has not been widely studied in Brazil.

Studies evaluating the timing of KRT in AKI have yielded conflicting results. Randomized trials in which the decision to initiate KRT was based on a creatinine cutoff point failed to show any mortality benefit of earlier KRT,^19-22^ whereas one in which the process of deciding to initiate KRT also incorporated other parameters of clinical deterioration showed that earlier KRT provided such a benefit.^23^ Additional analysis of a trial that failed to show a benefit of earlier KRT when the timing of its initiation was determined by the creatinine level revealed that there was in fact a detrimental effect if KRT was initiated too late,^24^ which is in accordance with the findings of previous observational studies.^25^ In a previous study, we also showed that early, daily hemodialysis improves survival among patients with Weil Syndrome (the severe form of leptospirosis).^26^

In this study, we aim to address the timing of KRT initiation, comparing the use of a temporal criterion with that of a serum creatinine cutoff criterion in terms of their association with in-hospital mortality in a population of critically ill patients with COVID-19-related AKI.

## Methods

### Study Design and Setting

This was a single-center, retrospective observational study conducted at the *Hospital das Clínicas* in São Paulo, Brazil, which is the largest city in Latin America, with an estimated population of more than 10 million (nearly 6% of the Brazilian population).^27^ In March 2020, the hospital became one of the referral centers for COVID-19 in the city of São Paulo. It is a quaternary referral center in general and has 900 beds, all of which were reserved exclusively for patients with moderate or severe COVID-19 during the first wave of the pandemic.

### Participants

All consecutive adult patients with COVID-19 and AKI who were admitted to the ICU between March 2020 to January 2021 were eligible for inclusion in the study if they were followed by nephrologists. All of the selected patients had laboratory-confirmed SARS-CoV-2 infection and AKI. Laboratory confirmation of SARS-CoV-2 infection was defined as a positive real-time reverse transcriptase-polymerase chain reaction assay of nasopharyngeal aspirates, nasal/throat swabs, or bronchial aspirate, or as positive serology for immunoglobulin M or G antibodies against SARS-CoV-2 in plasma. The diagnosis of AKI was based on the creatinine level and the need for KRT, in accordance with the Kidney Disease Improving Global Outcomes (KDIGO) criteria.^28^

Patients with stage 5 chronic kidney disease were excluded, as were those who were assigned to palliative care at admission, those who were admitted for a reason other than COVID-19, those who were kidney transplant patients, those in whom a diagnosis of COVID-19 had not been confirmed, those who were not admitted to the ICU, and those in whom KRT was initiated prior to admission. The study protocol was approved by the Institutional Review Board of the *Hospital das Clínicas* (Reference no. 4.129.320). Because of the retrospective nature of the study, the requirement for informed consent was waived.

### Data Sources

Baseline demographic data, including age and sex, were retrieved from electronic medical records, as were the following clinical and biochemical data: history of comorbidities; urine output; need for mechanical ventilation; Glasgow Coma Scale score; Richmond Agitation Sedation Scale score; vasopressor use; inotrope administration; blood pressure; partial pressure of arterial oxygen/fraction of inspired oxygen ratio; serum creatinine; platelet count; and total bilirubin. The Sequential Organ Failure Assessment (SOFA) score was also calculated.

The diagnosis of chronic kidney disease was based on the glomerular filtration rate, as estimated with the Chronic Kidney Disease-Epidemiology Collaboration equation,^29,30^ any value below 60 ml/min/1.73m^2^ being considered diagnostic. If there was no reliable baseline serum creatinine level on record, we used the lowest value measured during hospitalization without KRT or at least a week after KRT interruption.

The choice between intermittent hemodialysis and continuous venovenous hemodialysis (CVVHD) as the initial KRT modality was made by the attending nephrologist, on the basis of the clinical and hemodynamic characteristics of each patient. All analyses were based on the results of laboratory tests performed within the first 48 h of hospitalization or at the time of ICU admission.

### Potential Predictors

The main outcome measure was in-hospital mortality, the potential predictors of which were the timing of KRT initiation defined by a temporal criterion and that defined by a creatinine cutoff criterion. For the temporal criterion, patients were divided into two groups, those in whom KRT was initiated ≤ 6 days after admission (*n*=223) and those in whom it was initiated > 6 days after admission (*n*=169), designated the timely KRT and delayed KRT groups, respectively. The choice of 6 days as the cutoff was made because it was the median time between admission and start of KRT (Figure 1). For the creatinine cutoff criterion, patients were also divided into two groups, those in whom the serum creatinine level at KRT initiation was ≤ 4.74 mg/dl and those in whom it was > 4.74 mg/dl (4.74 mg/dl being the median value), each group comprising 196 patients. The cutoff at 4.74 was the median serum creatinine before the start of KRT. Within each criterion, the groups were analyzed in comparison with each other and in comparison with the no KRT group.

**Figure 1.**
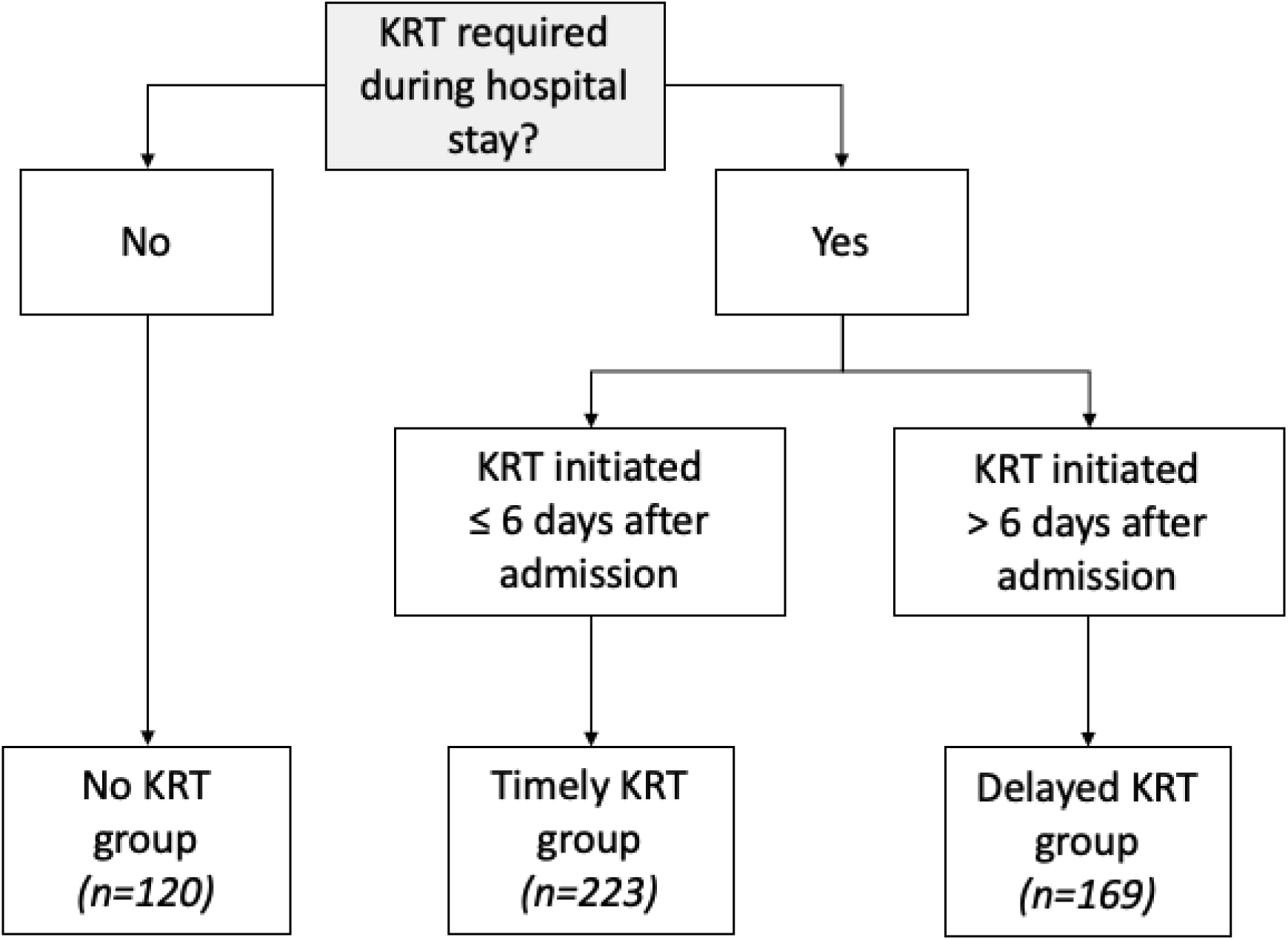
Composition of the groups evaluated in order to analyze the temporal criterion for determining the timing of kidney replacement therapy (KRT).

### Statistical Analysis

Continuous variables are reported as median and interquartile range (IQR), whereas categorical variables are reported as frequencies and percentages. For continuous variables, the differences among groups were evaluated with analysis of variance or with the Kruskal-Wallis test, as appropriate. Normality was tested by applying the Shapiro-Wilk test. For categorical variables, differences between groups were quantified with the chi-square test or Fisher’s exact test.

To evaluate the association between the timing of KRT initiation (by the temporal and creatinine cutoff criteria) and in-hospital mortality, we performed logistic regression, adjusting for other variables of interest. The other variables of interest were selected on the basis of previous reports of their association with in-hospital mortality. The results are displayed as odds ratios (ORs) and 95% confidence intervals (CIs). Selection of the best model was based on the Akaike information criterion. The level of statistical significance was set at *P* < 0.05.

For the purpose of calculating the SOFA score for multivariate logistic regression, simple imputation of the median value was used for missing bilirubin values, in 13 cases, and missing urine output values, in five cases. None of the other covariates retained in the final models required imputation. The prognostic performance of KRT timing was quantified by constructing a receiver operating characteristic and calculating the area under the curve (AUC).

Statistical analyses were performed in R software (version 4.1.3; R Foundation for Statistical Computing, Vienna, Austria), with the packages compareGroups (version 4.5.1), Hmisc (version 4.7-0), rms (version 6.3-0), and pROC (version 1.18.2).

## Results

### Characteristics of the Study Sample

Between March 2020 and January 2021, a total of 722 patients were followed by nephrologists at the *Hospital das Clínicas*. A total of 210 patients were excluded (Figure 2). Therefore, the final sample comprised 512 patients admitted to the ICU as a consequence of severe COVID-19.

**Figure 2.**
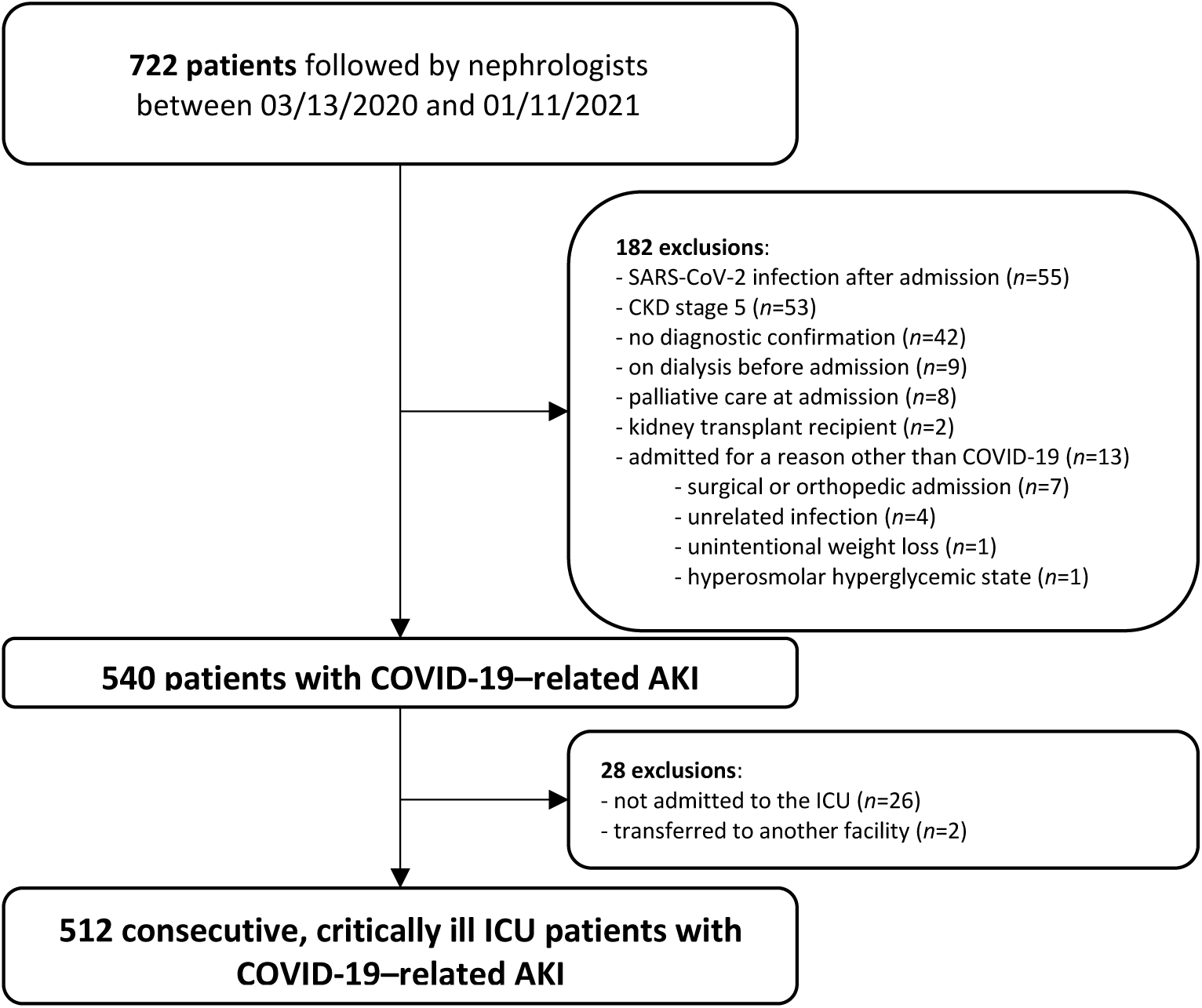
Flowchart of the patient selection process. SARS-CoV-2, severe acute respiratory syndrome coronavirus 2; CKD, chronic kidney disease, COVID-19, coronavirus disease 2019; AKI, acute kidney injury; ICU, intensive care unit.

The median (IQR) age was 64 (54-72) years, and 354 (69.1%) of the patients were men. Of the 512 patients evaluated, 335 (65.4%) had hypertension, 221 (43.2%) had diabetes mellitus, and 118 (23.0%) had cardiovascular disease; The median baseline serum creatinine was 1.07 mg/dl. At ICU admission, the median serum creatinine was 1.58 mg/dl and the median SOFA score was 10. At that time, 212 (41.4%) of the patients were receiving vasopressors, 361 (70.5%) were on mechanical ventilation, 367 (72.0%) were using corticosteroids, and 392 (76.6%) required KRT. In the sample as a whole, the in-hospital mortality rate was 72.5%. The median (IQR) time from hospital admission to the initiation of KRT was 6 (3-10) days.

Table 1 displays the characteristics of the sample according to the timing of KRT when the temporal criterion was applied (timely versus delayed KRT groups, in comparison with each other and with the no KRT group). In the timely KRT group, the proportion of patients with diabetes was higher; the SOFA scores at ICU admission were higher; the proportion of patients requiring vasopressor use was higher, as was that of those requiring mechanical ventilation; potassium levels were higher; pH values were lower; and serum creatinine levels at KRT initiation were higher. The baseline serum creatinine levels were lower in the no KRT group than in the timely and delayed KRT groups. In-hospital mortality was highest in the delayed KRT group. As the initial KRT modality, intermittent hemodialysis was implemented in 69.5% of the patients in the timely KRT group, whereas it was implemented in 52.1% of the patients in the late KRT group.

**Table 1.**
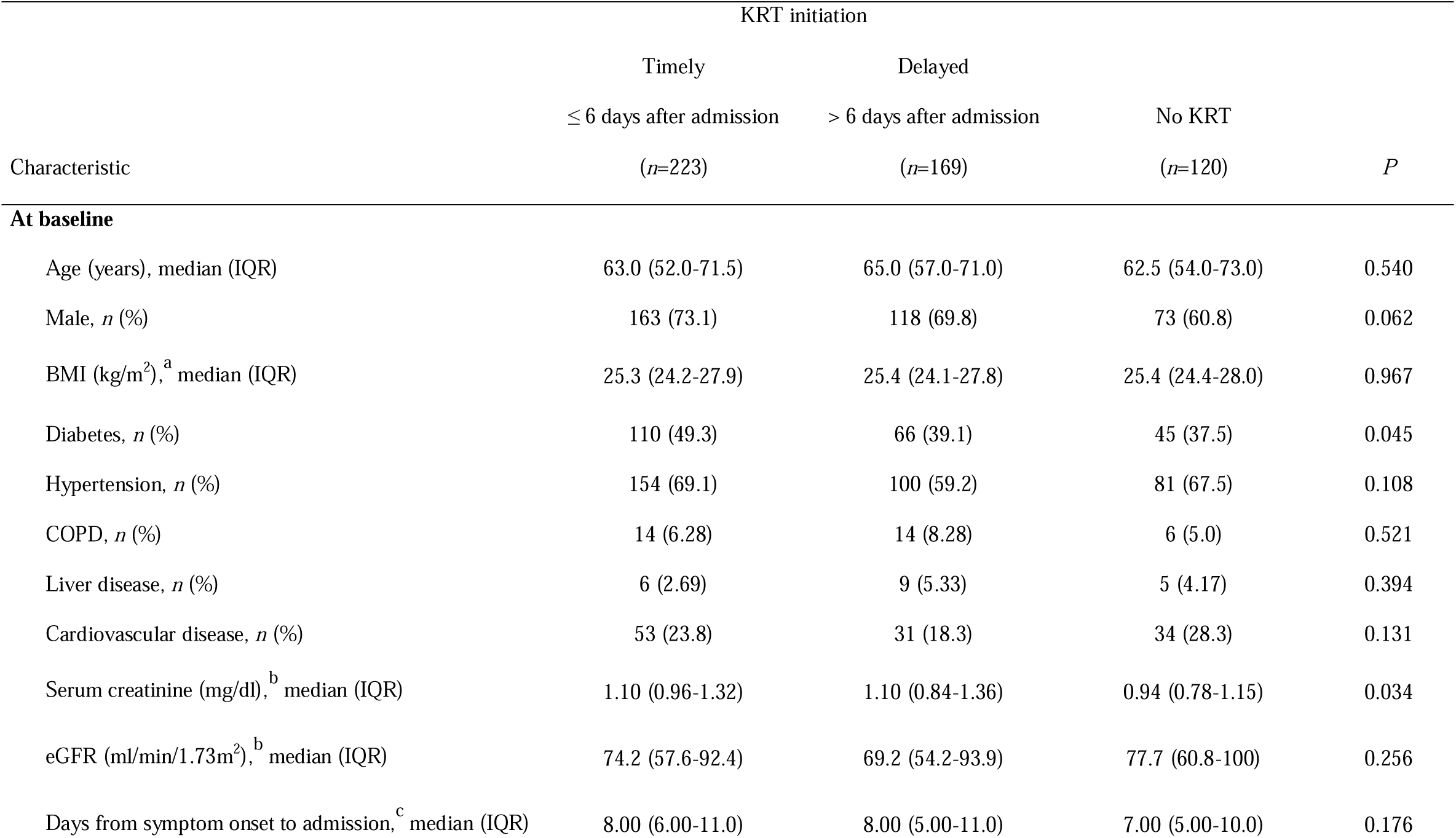
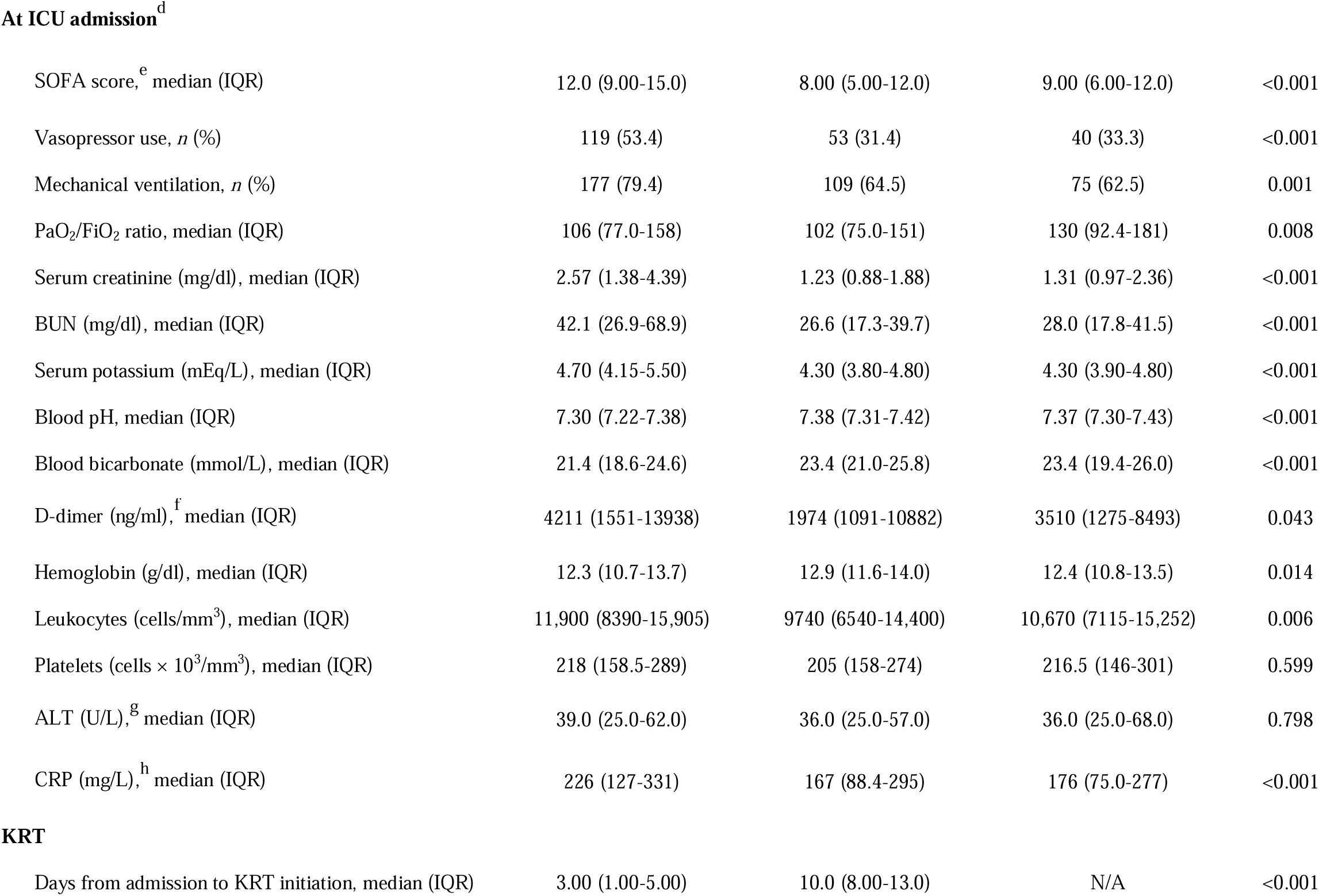
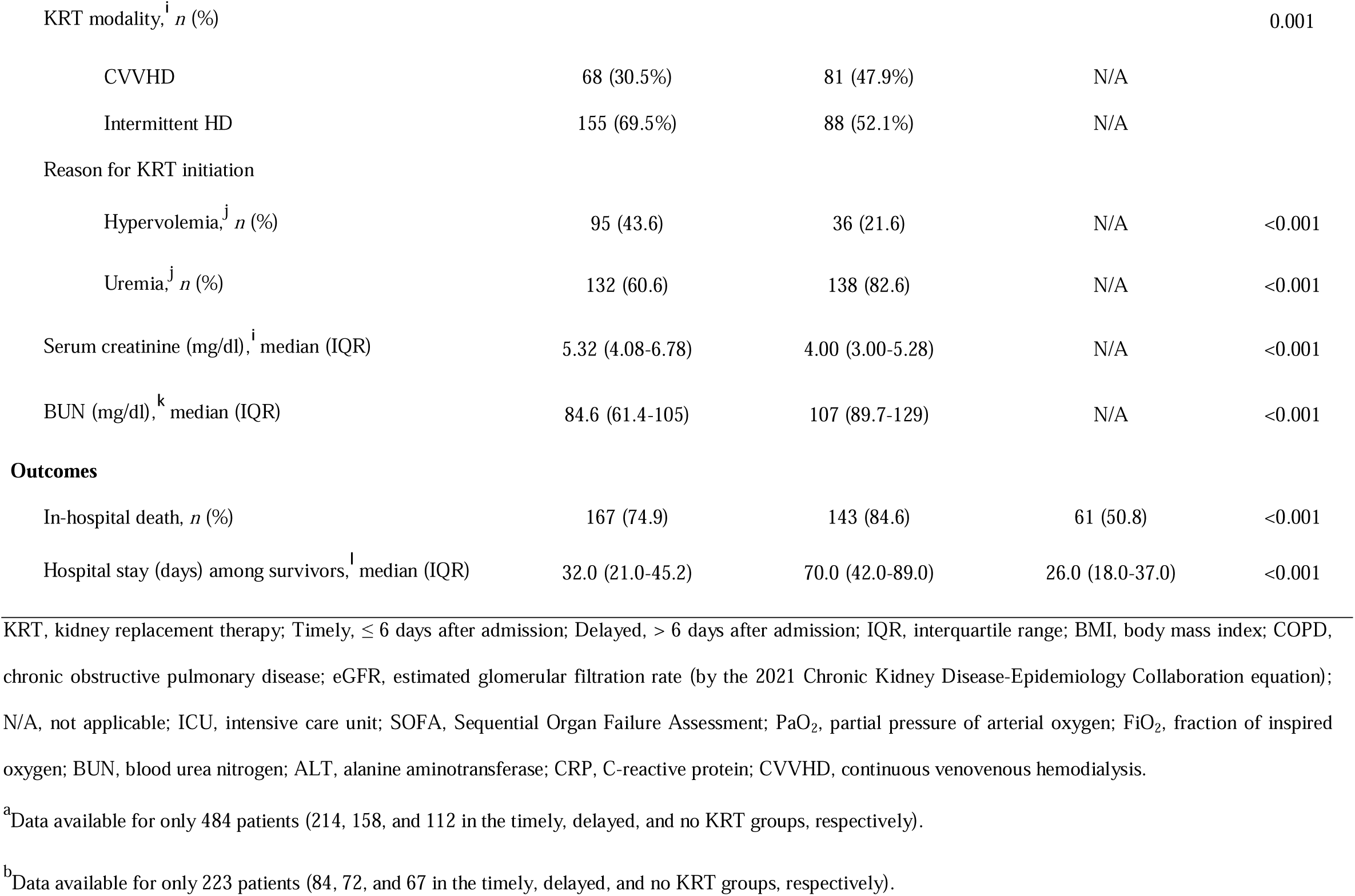
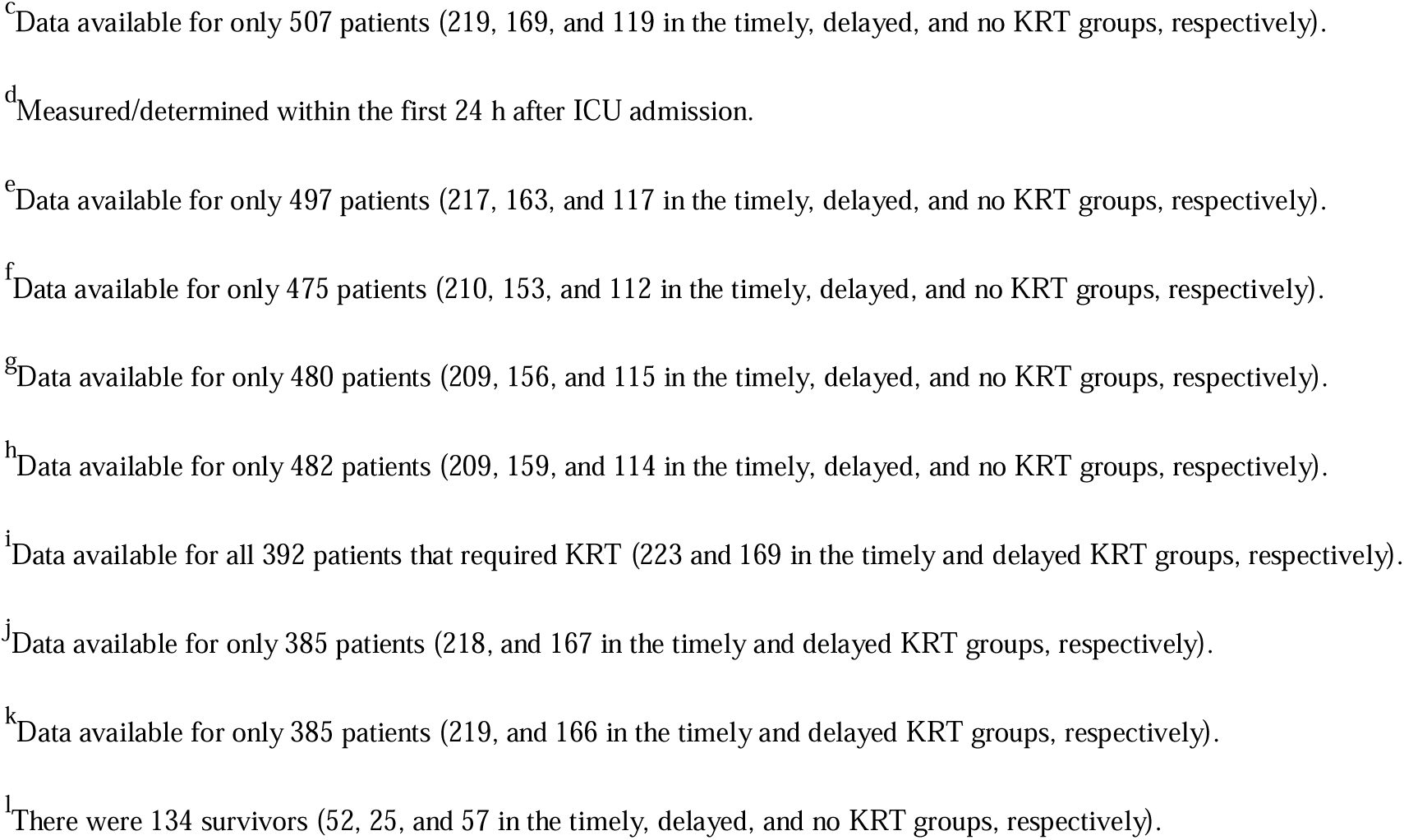
Characteristics of patients with coronavirus disease 2019-related acute kidney injury, by the timing of kidney replacement therapy, as determined on the basis of a temporal criterion.

Supplemental Table 1 shows the characteristics of the cohort stratified by the timing of KRT based on the serum creatinine level at KRT initiation. Supplemental Table 2 shows the characteristics of the cohort stratified by the initial KRT modality. In-hospital mortality was significantly higher among the patients in whom the initial modality was CVVHD than among those in whom it was intermittent hemodialysis (*P*=0.003).

### Timing of KRT and Prediction of In-Hospital Mortality

The logistic regression analyses for the temporal and creatinine cutoff criteria are summarized in Tables 2 and 3, respectively. In the unadjusted analyses for the temporal criterion (Table 2), delayed KRT was associated with a greater risk of in-hospital mortality (OR=1.84, vs. OR=0.35 for not requiring KRT). That association persisted after adjustment for age, sex, and SOFA score (model 1) or for age, sex, the need for mechanical ventilation, and vasopressor use (model 2). Likewise, not requiring KRT was associated with a lower risk of in-hospital mortality when compared with low serum creatinine (Table 3). However, high serum creatinine at KRT initiation was not associated with in-hospital mortality. That pattern persisted after adjustment for covariates (models 1 and 2).

**Table 2.**
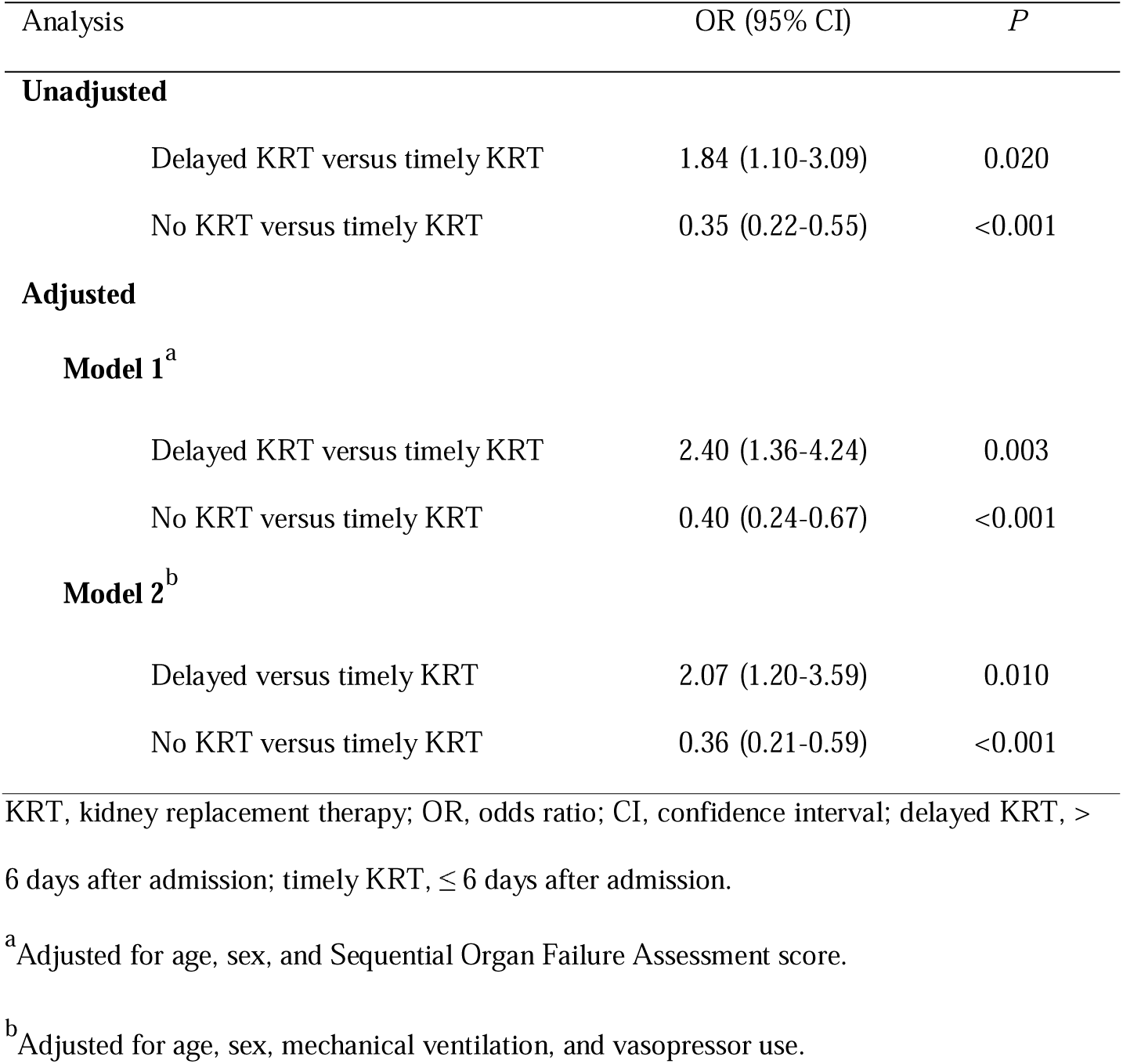
Logistic regression analyses of the association between the timing of kidney replacement therapy, as determined on the basis of a temporal criterion, and in-hospital mortality among patients with coronavirus disease 2019-related acute kidney injury.

| Analysis | OR (95% CI) | *P* |
| --- | --- | --- |
| **Unadjusted** |  |  |
| Delayed KRT versus timely KRT | 1.84 (1.10-3.09) | 0.020 |
| No KRT versus timely KRT | 0.35 (0.22-0.55) | <0.001 |
| **Adjusted** |  |  |
| **Model 1<sup>a</sup>** |  |  |
| Delayed KRT versus timely KRT | 2.40 (1.36-4.24) | 0.003 |
| No KRT versus timely KRT | 0.40 (0.24-0.67) | <0.001 |
| **Model 2<sup>b</sup>** |  |  |
| Delayed versus timely KRT | 2.07 (1.20-3.59) | 0.010 |
| No KRT versus timely KRT | 0.36 (0.21-0.59) | <0.001 |

KRT, kidney replacement therapy; OR, odds ratio; CI, confidence interval; delayed KRT, > 6 days after admission; timely KRT, ≤ 6 days after admission.
<sup>a</sup>Adjusted for age, sex, and Sequential Organ Failure Assessment score.
<sup>b</sup>Adjusted for age, sex, mechanical ventilation, and vasopressor use.

**Table 3.** Logistic regression analyses of the association between the timing of kidney replacement therapy, as determined on the basis of a serum creatinine cutoff criterion, and in-hospital mortality among patients with coronavirus disease 2019-related acute kidney injury.

| Analysis | OR (95% CI) | *P* |
| --- | --- | --- |
| **Unadjusted** |  |  |
| High sCr versus low sCr | 0.65 (0.40-1.06) | 0.083 |
| No KRT versus low sCr | 0.22 (0.13-0.36) | <0.001 |
| **Adjusted** |  |  |
| **Model 1<sup>a</sup>** |  |  |
| High sCr versus low sCr | 0.60 (0.35-1.03) | 0.062 |
| No KRT versus low sCr | 0.21 (0.12-0.36) | <0.001 |
| **Model 2<sup>b</sup>** |  |  |
| High sCr versus low sCr | 0.65 (0.38-1.10) | 0.110 |
| No KRT versus low sCr | 0.21 (0.12-0.36) | <0.001 |

KRT, kidney replacement therapy; OR, odds ratio; CI, confidence interval; high sCr, serum creatinine > 4.74 mg/dl at KRT initiation; low sCr, serum creatinine ≤ 4.74 mg/dl at KRT initiation.
<sup>a</sup>Adjusted for age, sex, and Sequential Organ Failure Assessment score.
<sup>b</sup>Adjusted for age, sex, mechanical ventilation, and vasopressor use.

Figure 3 depicts the relationship between in-hospital mortality and the timing of KRT initiation when the temporal criterion was applied. Longer delays were associated with higher in-hospital mortality rates (*P* < 0.001).

**Figure 3.**
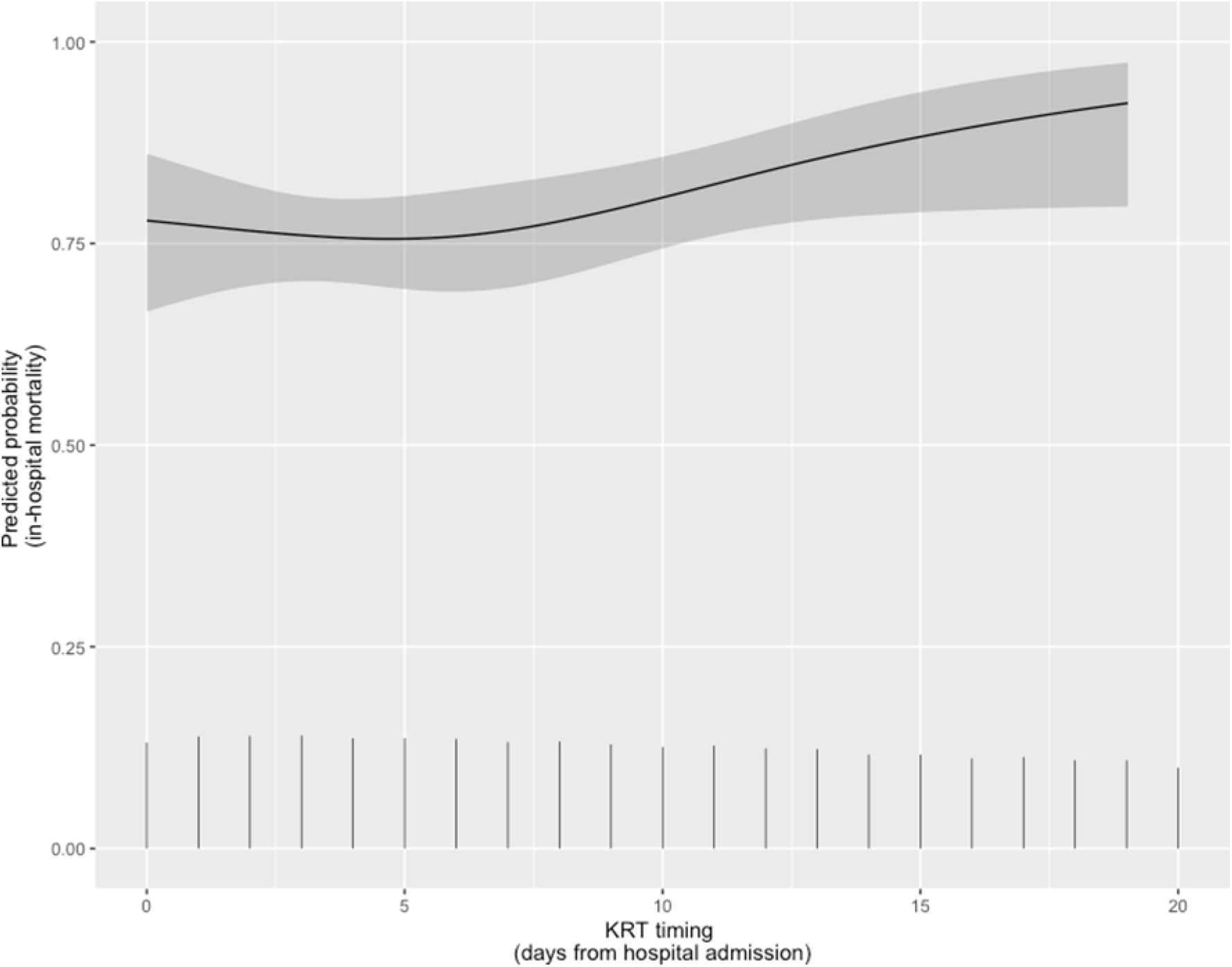
Restricted cubic spline model depicting the unadjusted relationship (with 95% confidence intervals) between the timing of kidney replacement therapy (KRT), as determined on the basis of a temporal criterion, and the predicted probability of in-hospital mortality in patients with coronavirus disease 2019-related acute kidney injury. Vertical lines show the spike histogram.

### Performance of KRT Timing for the Prediction of In-Hospital Mortality

Table 4 shows the AUCs for the prediction of in-hospital mortality for KRT timing by the temporal and creatinine cutoff criteria. Both criteria performed similarly, with AUCs of 0.662 and 0.653, respectively.

**Table 4.** Receiver operating characteristic curve analyses of the performance of timing of kidney replacement therapy for the prediction of in-hospital mortality among patients with coronavirus disease 2019-related acute kidney injury.

| Analysis | AUC (95% CI) |
| --- | --- |
| KRT timing overall, in model 1<sup>a</sup> | 0.698 (0.647-0.749) |
| KRT timing based on the temporal criterion (unadjusted) | 0.662 (0.612-0.712) |
| KRT timing based on the temporal criterion, in model 1<sup>a</sup> | 0.767 (0.720-0.813) |
| KRT timing based on the temporal criterion, in model 2<sup>b</sup> | 0.760 (0.713-0.807) |
| KRT timing based on the serum creatinine cutoff criterion (unadjusted) | 0.653 (0.602-0.705) |
| KRT timing based on the serum creatinine cutoff criterion, in model 1<sup>a</sup> | 0.754 (0.707-0.802) |
| KRT timing based on the serum creatinine cutoff criterion, in model 2<sup>b</sup> | 0.752 (0.704-0.800) |

AUC, area under the curve; CI, confidence interval; KRT, kidney replacement therapy.
<sup>a</sup> Adjusted for age, sex, and Sequential Organ Failure Assessment score.
<sup>b</sup> Adjusted for age, sex, mechanical ventilation, and vasopressor use.

## Discussion

Although some of the studies conducted at the beginning of the pandemic showed that kidney involvement was not very common in patients with COVID-19,^16,17^ we now know that kidney disease at admission and the development of AKI during hospitalization are highly prevalent in patients with COVID-19 and are associated with in-hospital mortality.^31-36^ Mortality rates of up to 70% have been reported among patients with COVID-19-related AKI.^37^ The few studies of the topic in Brazil have reported mortality rates ranging from 72% to 93%.^26,27^ In the present study, the overall mortality rate was 72.6%, which is comparable to the rates reported elsewhere.

The timing of KRT in patients with COVID-19-related AKI and its impact on clinical outcomes have not been widely described. In non-urgent cases of severe AKI (such as those related to refractory hyperkalemia, metabolic acidosis, or volume overload), the role of KRT is less clear, as discussed in a recent review of the literature.^38^

In one randomized clinical trial of surgical patients with KDIGO stage 2 AKI, with a neutrophil gelatinase-associated lipocalin level > 150 ng/ml and at least one comorbidity (e.g., sepsis, vasopressor use, and volume overload), the 90-day mortality rate was 39% among the patients receiving early KRT, versus 54% among those receiving delayed KRT.^32^ Another, multicenter, randomized trial of patients with KDIGO stage 3 AKI who were on mechanical ventilation, treated or not with catecholamines, and assigned to an early or a delayed strategy of KRT showed that 60-day mortality was comparable between the two groups (48.5% versus 49.7%).^28^ In that study, some of the patients in the delayed-strategy group did not need dialysis. However, among the patients who received delayed KRT, 60-day mortality was 61.8%, significantly higher than the 48.5% reported for the early-strategy group. A subsequent multicenter, open-label, randomized, controlled trial, conducted by the same group, showed that, in patients with AKI who had oliguria for more than 72 h or a blood urea nitrogen concentration higher than 112 mg/dl and no severe complication that would warrant immediate KRT, a longer delay before the initiation of KRT did not confer additional benefit.^33^ Although 767 patients were randomized in that study, 127 received KRT because of an urgent indication (before meeting the randomization criteria); an additional 352 did not meet the randomization criteria and therefore did not receive KRT. Consequently, the final sample comprised 278 patients. The initial KRT modality was intermittent dialysis in approximately 60% of those patients. The authors demonstrated that although most of the patients were receiving vasoactive drugs, they were able to tolerate the intermittent dialysis. In a multinational, randomized, controlled trial involving patients with KDIGO stage 2 or 3 AKI, the 90-day mortality rate did not differ significantly between the patients in whom a standard KRT strategy was used and those in whom an accelerated KRT strategy was used.^29^ In the present study, we found that, if the timing of KRT was defined on the basis of the temporal criterion, the in-hospital mortality rate among patients with COVID-19-related AKI was significantly higher when KRT was delayed. When the timing of KRT was defined on the basis of the creatinine cutoff criterion, we found no such difference.

It is questionable whether monitoring serum creatinine values and conducting regular follow-up examinations are sufficient to predict AKI and the need for KRT. As discussed in one recent study,^39^ AKI should be thought of not as a single disease but rather as a condition provoked by a collection of syndromes such as urinary tract obstruction, cardiorenal syndrome, and sepsis, as well as the more current COVID-19. According to the authors of that study, the approach to a patient with AKI should depend on the clinical context and can also vary depending on resource availability, which was a challenge during the COVID-19 pandemic. Many factors can contribute to changes in creatinine, which led us to think about what else could be added to the assessment of critically ill patients.^40,41^ Serum creatinine is far from being the best indicator of kidney function. It has also been known that creatinine is of limited use as a marker of kidney injury in sepsis, in which its production is reduced.^42^

In the present study, a high SOFA score was found to correlate with the need for KRT. That might facilitate the decision of whether or not to initiate KRT in a critically ill patient. Most of the studies conducted to date have analyzed only the risk factors for COVID-19-related AKI, without analyzing the compound factors evaluated here.

Critically ill patients with COVID-19 usually develop severe respiratory insufficiency, requiring a high fraction of inspired oxygen. In such patients, fluid accumulation has been associated with adverse outcomes.^43,44^ Therefore, initiating KRT earlier and avoiding fluid overload might improve survival among patients with COVID-19. Some studies have also shown that, for patients with COVID-19-related AKI, early initiation of KRT or another form of extracorporeal organ support seems to prevent progression of the disease.^45-48^ In addition, the decision to initiate KRT should be individualized and should take into consideration the current demand and capacity.^49-51^

Timely initiation of KRT seems to be extremely important in the context of COVID-19-related AKI in the ICU. In the present study, we evaluated KRT timing in a group of patients with the same disease, although with various comorbidities. We found that timely KRT initiation reduced ICU mortality. To our knowledge, this is the first study to report the timing of KRT during the COVID-19 pandemic.

Our study has some limitations. The single-center design likely reduced the effect size. In addition, because it was an observational study conducted in the setting of the COVID-19 pandemic, causal relationships cannot be inferred from our results. The division of groups by time is justified above, and we know that the pandemic scenario may have been a defining factor in having a group starting late (> 6 days). Furthermore, the fact that we analyzed data only for cases followed by nephrologists could have created a selection bias, because intensivists felt confident to manage mild cases of AKI and because severe cases of AKI were considered unlikely to benefit from nephrology consultation or KRT.

## Conclusion

In ICU patients with COVID-19-related AKI, the in-hospital mortality rate is high. Among such patients, earlier initiation of KRT seems to be associated with lower in-hospital mortality when a temporal criterion is applied.

## Data Availability

All data produced in the present study are available upon reasonable request to the authors

## Disclosures

C.A.P. de Almeida is a member of the Global Medical Review team at Eli Lilly and Company. P.R.G.L. started working at Baxter Brasil in June/2023. V.F.S. received a consulting fee for participation on an Advisory Board meeting in March/2022 for Vifor Pharma Brazil, outside of the submitted work. C.E.R. received fees from Medtronic for providing instruction in catheter insertion in 2021. The remaining authors have nothing to disclose.

## Funding

This work was supported by the *Fundação de Amparo à Pesquisa do Estado de São Paulo* (FAPESP, São Paulo Research Foundation; Grant no. 2020/06583-1). L. Andrade is the recipient of a grant from the Brazilian *Conselho Nacional de Desenvolvimento Científico e Tecnológico* (National Council for Scientific and Technological Development; Grant no. 309683/2021-1).

## Author Contributions

**Conceptualization:** C.A.P. de Almeida, M.F.A. de Oliveira, L. Andrade.

**Data curation:** C.A.P. de Almeida, M.F.A. de Oliveira, A.M. Teixeira, C.P.S. Cabrera, I. Smolentzov, P.R.G. Lins, C.E. Rodrigues, V.F. Seabra, L. Andrade.

**Formal analysis:** C.A.P. de Almeida, V.F. Seabra, L. Andrade.

**Methodology:** C.A.P. de Almeida, M.F.A. de Oliveira, A.M. Teixeira, C.P.S. Cabrera, I. Smolentzov, B.V. Reichert, P.R.G. Lins, C.E. Rodrigues, V.F. Seabra, L. Andrade.

**Project administration:** C.A.P. de Almeida, M.F.A. de Oliveira, L. Andrade.

**Supervision:** L. Andrade.

**Writing - original draft:** C.A.P. de Almeida, V.F. Seabra, L. Andrade.

**Writing - review & editing:** C.A.P. de Almeida, M.F.A. de Oliveira, A.M. Teixeira, C.P.S. Cabrera, I. Smolentzov, B.V. Reichert, P.R.G. Lins, C.E. Rodrigues, V.F. Seabra, L. Andrade.

## Data Sharing Statement

The data employed in this work are available from the corresponding author on reasonable request.

## Notes

### Competing Interest Statement

The authors have declared no competing interest.

### Funding Statement

This work was supported by the Sao Paulo Research Foundation (FAPESP); Grant no. 2020/06583-1). L. Andrade is the recipient of a grant from the Brazilian National Council for Scientific and Technological Development (CNPq; Grant no. 309683/2021-1).

### Author Declarations

The study protocol was approved by the Institutional Review Board of the Hospital das Clinicas (Reference no. 4.129.320). Because of the retrospective nature of the study, the requirement for informed consent was waived.

