## Supplemental Tables for "Kidney Replacement Therapy in COVID-19-Related Acute Kidney Injury: the Importance of Timing"

**Supplemental Table 1. Characteristics of patients with coronavirus disease 2019-related acute kidney injury, by the timing of kidney replacement therapy, as defined by a temporal criterion**

| Characteristic | KRT initiation | | No KRT | *P* |
| --- | --- | --- | --- | --- |
|  | Earlier | Delayed |  |  |
|  | (≤ 6 days after admission) | (> 6 days after admission) |  |  |
|  | (*n*=223) | (*n*=169) | (*n*=120) |  |
| **At baseline** |  |  |  |  |
| Age (years), median (IQR) | 63.0 (52.0–71.5) | 65.0 (57.0–71.0) | 62.5 (54.0–73.0) | 0.540 |
| Male, *n* (%) | 163 (73.1) | 118 (69.8) | 73 (60.8) | 0.062 |
| BMI (kg/m^2^),^a^ median (IQR) | 25.3 (24.2–27.9) | 25.4 (24.1–27.8) | 25.4 (24.4–28.0) | 0.967 |
| Diabetes, *n* (%) | 110 (49.3) | 66 (39.1) | 45 (37.5) | 0.045 |
| Hypertension, *n* (%) | 154 (69.1) | 100 (59.2) | 81 (67.5) | 0.108 |
| COPD, *n* (%) | 14 (6.28) | 14 (8.28) | 6 (5.0) | 0.521 |
| Liver disease, *n* (%) | 6 (2.69) | 9 (5.33) | 5 (4.17) | 0.394 |
| Cardiovascular disease, *n* (%) | 53 (23.8) | 31 (18.3) | 34 (28.3) | 0.131 |
| Serum creatinine (mg/dl),^b^ median (IQR) | 1.10 (0.96–1.32) | 1.10 (0.84–1.36) | 0.94 (0.78–1.15) | 0.034 |
| eGFR (ml/min/1.73m^2^),^b^ median (IQR) | 74.2 (57.6–92.4) | 69.2 (54.2–93.9) | 77.7 (60.8–100) | 0.256 |
| Days from symptom onset to admission,^c^ median (IQR) | 8.00 (6.00–11.0) | 8.00 (5.00–11.0) | 7.00 (5.00–10.0) | 0.176 |
| **At ICU admission**^d^ |  |  |  |  |
| SOFA score,^e^ median (IQR) | 12.0 (9.00–15.0) | 8.00 (5.00–12.0) | 9.00 (6.00–12.0) | <0.001 |
| Vasopressor use, *n* (%) | 119 (53.4) | 53 (31.4) | 40 (33.3) | <0.001 |
| Mechanical ventilation, *n* (%) | 177 (79.4) | 109 (64.5) | 75 (62.5) | 0.001 |
| PaO_2_/FiO_2_ ratio, median (IQR) | 106 (77.0–158) | 102 (75.0–151) | 130 (92.4–181) | 0.008 |
| Serum creatinine (mg/dl), median (IQR) | 2.57 (1.38–4.39) | 1.23 (0.88–1.88) | 1.31 (0.97–2.36) | <0.001 |
| BUN (mg/dl), median (IQR) | 42.1 (26.9–68.9) | 26.6 (17.3–39.7) | 28.0 (17.8–41.5) | <0.001 |
| Serum potassium (mEq/L), median (IQR) | 4.70 (4.15–5.50) | 4.30 (3.80–4.80) | 4.30 (3.90–4.80) | <0.001 |
| Blood pH, median (IQR) | 7.30 (7.22–7.38) | 7.38 (7.31–7.42) | 7.37 (7.30–7.43) | <0.001 |
| Blood bicarbonate (mmol/L), median (IQR) | 21.4 (18.6–24.6) | 23.4 (21.0–25.8) | 23.4 (19.4–26.0) | <0.001 |
| D-dimer (ng/ml),^f^ median (IQR) | 4211 (1551–13938) | 1974 (1091–10882) | 3510 (1275–8493) | 0.043 |
| Hemoglobin (g/dl), median (IQR) | 12.3 (10.7–13.7) | 12.9 (11.6–14.0) | 12.4 (10.8–13.5) | 0.014 |
| Leukocytes (cells/mm^3^), median (IQR) | 11,900 (8390–15,905) | 9740 (6540–14,400) | 10,670 (7115–15,252) | 0.006 |
| Platelets (cells × 10^3^/mm^3^), median (IQR) | 218 (158.5–289) | 205 (158–274) | 216.5 (146–301) | 0.599 |
| ALT (U/L),^g^ median (IQR) | 39.0 (25.0–62.0) | 36.0 (25.0–57.0) | 36.0 (25.0–68.0) | 0.798 |
| CRP (mg/L),^h^ median (IQR) | 226 (127–331) | 167 (88.4–295) | 176 (75.0–277) | <0.001 |
| **KRT** |  |  |  |  |
| Days from admission to KRT initiation, median (IQR) | 3.00 (1.00–5.00) | 10.0 (8.00–13.0) | N/A | <0.001 |
| KRT modality,^i^ *n* (%) |  |  |  | 0.001 |
| CVVHD | 68 (30.5%) | 81 (47.9%) | N/A |  |
| Intermittent HD | 155 (69.5%) | 88 (52.1%) | N/A |  |
| Reason for KRT initiation |  |  |  |  |
| Hypervolemia,^j^ *n* (%) | 95 (43.6) | 36 (21.6) | N/A | <0.001 |
| Uremia,^j^ *n* (%) | 132 (60.6) | 138 (82.6) | N/A | <0.001 |
| Serum creatinine (mg/dl),^i^ median (IQR) | 5.32 (4.08–6.78) | 4.00 (3.00–5.28) | N/A | <0.001 |
| BUN (mg/dl),^k^ median (IQR) | 84.6 (61.4–105) | 107 (89.7–129) | N/A | <0.001 |
| **Outcomes** |  |  |  |  |
| In-hospital death, *n* (%) | 167 (74.9) | 143 (84.6) | 61 (50.8) | <0.001 |
| Hospital stay (days) among survivors,^l^ median (IQR) | 32.0 (21.0–45.2) | 70.0 (42.0–89.0) | 26.0 (18.0–37.0) | <0.001 |

KRT, kidney replacement therapy; IQR, interquartile range; BMI, body mass index; COPD, chronic obstructive pulmonary disease; eGFR, estimated glomerular filtration rate (by the 2021 Chronic Kidney Disease–Epidemiology Collaboration equation); N/A, not applicable; ICU, intensive care unit; SOFA, Sequential Organ Failure Assessment; PaO_2_, partial pressure of arterial oxygen; FiO_2_, fraction of inspired oxygen; BUN, blood urea nitrogen; ALT, alanine aminotransferase; CRP, C-reactive protein; CVVHD, continuous venovenous hemodialysis.

^a^Data available for only 484 patients (214, 158, and 112 in the earlier, later, and no KRT groups, respectively).

^b^Data available for only 223 patients (84, 72, and 67 in the earlier, later, and no KRT groups, respectively).

^c^Data available for only 507 patients (219, 169, and 119 in the earlier, later, and no KRT groups, respectively).

^d^Measured/determined within the first 24 h after ICU admission.

^e^Data available for only 497 patients (217, 163, and 117 in the earlier, later, and no KRT groups, respectively).

^f^Data available for only 475 patients (210, 153, and 112 in the earlier, later, and no KRT groups, respectively).

^g^Data available for only 480 patients (209, 156, and 115 in the earlier, later, and no KRT groups, respectively).

^h^Data available for only 482 patients (209, 159, and later114 in the earlier, later, and no KRT groups, respectively).

^i^Data available for all 392 patients that required KRT (223 and 169 in the earlier and later KRT groups, respectively).

^j^Data available for only 385 patients (218, and 167 in the earlier and later KRT groups, respectively).

^k^Data available for only 385 patients (219, and 166 in the earlier and later KRT groups, respectively).

^l^There were 134 survivors (52, 25, and 57 in the earlier, later, and no KRT groups, respectively).

**Supplemental Table 2. Characteristics of patients with coronavirus disease 2019-related acute kidney injury, by the timing of kidney replacement therapy, as defined by a serum creatinine cutoff criterion.**

| Characteristic | Serum creatinine at KRT initiation | | No KRT | *P* |
| --- | --- | --- | --- | --- |
|  | Low | High |  |  |
|  | (≤ 4.74 mg/dl) | (> 4.74 mg/dl) |  |  |
|  | (*n*=196) | (*n*=196) | (*n*=120) |  |
| **At baseline** |  |  |  |  |
| Age (years), median (IQR) | 65.0 (57.0–71.0) | 63.0 (52.0–71.0) | 62.5 (54.0–73.0) | 0.234 |
| Male, *n* (%) | 122 (62.2) | 159 (81.1) | 73 (60.8) | <0.001 |
| BMI (kg/m^2^),^a^ median (IQR) | 25.4 (24.2–28.1) | 25.3 (24.1–27.6) | 25.4 (24.4–28.0) | 0.45 |
| Diabetes, *n* (%) | 87 (44.4) | 89 (45.4) | 45 (37.5) | 0.351 |
| Hypertension, *n* (%) | 113 (57.7) | 141 (71.9) | 81 (67.5) | 0.01 |
| COPD, *n* (%) | 19 (9.69) | 9 (4.59) | 6 (5.0) | 0.091 |
| Liver disease, *n* (%) | 9 (4.59) | 6 (3.06) | 5 (4.17) | 0.771 |
| Cardiovascular disease, *n* (%) | 40 (20.4) | 44 (22.4) | 34 (28.3) | 0.259 |
| Serum creatinine (mg/dl),^b^ median (IQR) | 1.10 (0.84–1.35) | 1.12 (0.96–1.30) | 0.94 (0.78–1.15) | 0.035 |
| eGFR (ml/min/1.73m^2^),^b^ median (IQR) | 68.9 (53.4–93.7) | 74.3 (61.2–92.4) | 77.7 (60.8–100) | 0.146 |
| Days from symptom onset to admission,^c^ median (IQR) | 8.00 (5.00–11.0) | 8.00 (6.00–11.0) | 7.00 (5.00–10.0) | 0.058 |
| Serum creatinine (mg/dl) at KRT initiation, median (IQR) | 3.51 (2.75–4.13) | 6.35 (5.37–7.27) | N/A | <0.001 |
| **At ICU admission**^d^ |  |  |  |  |
| SOFA score,^e^ median (IQR) | 9.00 (6.00–13.0) | 11.0 (8.00–14.0) | 9.00 (6.00–12.0) | <0.001 |
| Vasopressor use, *n* (%) | 89 (45.4) | 83 (42.3) | 40 (33.3) | 0.101 |
| Mechanical ventilation, *n* (%) | 135 (68.9) | 151 (77.0) | 75 (62.5) | 0.019 |
| PaO_2_/FiO_2_ ratio, median (IQR) | 107 (76.8–153) | 103 (77.0–156) | 130 (92.4–181) | 0.008 |
| Serum creatinine (mg/dl), median (IQR) | 1.51 (0.98–2.46) | 2.29 (1.23–4.82) | 1.31 (0.97–2.36) | <0.001 |
| BUN (mg/dl), median (IQR) | 31.8 (19.0–46.7) | 37.4 (24.8–64.8) | 28.0 (17.8–41.5) | <0.001 |
| Serum potassium (mEq/L), median (IQR) | 4.40 (3.90–5.00) | 4.60 (4.10–5.43) | 4.30 (3.90–4.80) | <0.001 |
| Blood pH, median (IQR) | 7.35 (7.26–7.41) | 7.32 (7.24–7.40) | 7.37 (7.30–7.43) | 0.009 |
| Blood bicarbonate (mmol/L), median (IQR) | 22.7 (19.6–25.4) | 22.4 (20.0–25.1) | 23.4 (19.4–26.0) | 0.384 |
| D-dimer (ng/ml),^f^ median (IQR) | 2193 (1224–6851) | 4303 (1320–15873) | 3510 (1275–8493) | 0.098 |
| Hemoglobin (g/dl), median (IQR) | 12.4 (11.1–13.8) | 12.7 (11.1–13.9) | 12.4 (10.8–13.5) | 0.137 |
| Leukocytes (cells/mm^3^), median (IQR) | 10,080 (6635–15,078) | 11,715 (8520–15,622) | 10,670 (7115–15,252) | 0.063 |
| Platelets (cells × 10^3^/mm^3^), median (IQR) | 204.5 (155.8–267.3) | 218.5 (167–296.5) | 216.5 (146–301) | 0.165 |
| ALT (U/L),^g^ median (IQR) | 36.5 (23.0–60.0) | 39.0 (27.0–57.5) | 36.0 (25.0–68.0) | 0.717 |
| CRP (mg/L),^h^ median (IQR) | 180 (96.0–298) | 220 (131–331) | 176 (75.0–277) | 0.003 |
| **KRT** |  |  |  |  |
| Days from admission to KRT initiation, median (IQR) | 7.00 (4.00–11.0) | 4.00 (2.00–7.00) | N/A | <0.001 |
| KRT modality,^i^ *n* (%) |  |  |  | 0.013 |
| CVVHD | 87 (44.4) | 62 (31.6) | N/A |  |
| Intermittent HD | 109 (55.6) | 134 (68.4) | N/A |  |
| Reason for KRT initiation |  |  |  |  |
| Hypervolemia,^j^ *n* (%) | 67 (34.9) | 64 (33.2) | N/A | 0.801 |
| Uremia,^j^ *n* (%) | 128 (66.7) | 142 (73.6) | N/A | 0.171 |
| Serum creatinine (mg/dl),^i^ median (IQR) | 3.51 (2.75–4.13) | 6.35 (5.37–7.27) | N/A | <0.001 |
| BUN (mg/dl),^j^ median (IQR) | 93.9 (61.2–113) | 96.7 (75.2–117) | N/A | 0.102 |
| **Outcomes** |  |  |  |  |
| In-hospital death, *n* (%) | 162 (82.7) | 148 (75.5) | 61 (50.8) | <0.001 |
| Hospital stay (days) among survivors,^k^ median (IQR) | 45.0 (32.0–73.0) | 33.5 (21.0–52.5) | 26.0 (18.0–37.0) | <0.001 |

KRT, kidney replacement therapy; IQR, interquartile range; BMI, body mass index; COPD, chronic obstructive pulmonary disease; eGFR, estimated glomerular filtration rate (by the 2021 Chronic Kidney Disease–Epidemiology Collaboration equation); N/A, not applicable; ICU, intensive care unit; SOFA, Sequential Organ Failure Assessment; PaO_2_, partial pressure of arterial oxygen; FiO_2_, fraction of inspired oxygen; BUN, blood urea nitrogen; ALT, alanine aminotransferase; CRP, C-reactive protein; CVVHD, continuous venovenous hemodialysis.

^a^Data available for only 484 patients (186, 186, and 112 in the low, high, and no KRT groups, respectively).

^b^Data available for only 223 patients (88, 68, and 67 in the low, high, and no KRT groups, respectively).

^c^Data available for only 507 patients (195, 193, and 119 in the low, high, and no KRT groups, respectively).

^d^Measured/determined within the first 24 h after ICU admission.

^e^Data available for only 497 patients (189, 191, and 117 in the low, high, and no KRT groups, respectively).

^f^Data available for only 475 patients (182, 181, and 112 in the low, high, and no KRT groups, respectively).

^g^Data available for only 480 patients (182, 183, and 115 in the low, high, and no KRT groups, respectively).

^h^Data available for only 482 patients (184, 184, and 114 in the low, high, and no KRT groups, respectively).

^i^Data available for all 392 patients that required KRT (196 and 196 in the low and high groups, respectively).

^j^Data available for only 385 patients (192 and 193 in the low and high groups, respectively).

^k^There were 134 survivors (33, 44, and 57 in the low, high, and no KRT groups, respectively).

**Supplemental Table 3. Characteristics of patients with coronavirus disease 2019-related acute kidney injury, by initial kidney replacement therapy modality.**

| Characteristic | CVVHD | Intermittent HD | *P* |
| --- | --- | --- | --- |
|  | (*n*=149) | (*n*=243) |  |
| **Baseline** |  |  |  |
| Age (years), median (IQR) | 65.0 (54.0–72.0) | 63.0 (53.0–71.0) | 0.353 |
| Male, *n* (%) | 109 (73.2) | 172 (70.8) | 0.696 |
| BMI (kg/m^2^),^a^ median (IQR) | 25.3 (24.2–27.5) | 25.4 (24.2–28.1) | 0.269 |
| Diabetes, *n* (%) | 64 (43.0) | 112 (46.1) | 0.616 |
| Hypertension, *n* (%) | 96 (64.4) | 158 (65.0) | 0.992 |
| COPD, *n* (%) | 12 (8.05) | 16 (6.58) | 0.729 |
| Liver disease, *n* (%) | 7 (4.7) | 8 (3.29) | 0.665 |
| Cardiovascular disease, *n* (%) | 38 (25.5) | 46 (18.9) | 0.158 |
| Serum creatinine (mg/dl),^b^ median (IQR) | 1.10 (0.97–1.31) | 1.10 (0.88–1.35) | 0.717 |
| eGFR (ml/min/1.73m^2^),^b^ median (IQR) | 68.6 (55.0–80.1) | 73.8 (56.5–95.6) | 0.221 |
| **At ICU admission**^c^ |  |  |  |
| SOFA score, median (IQR) | 11.0 (7.00–14.0) | 10.5 (7.00–13.0) | 0.359 |
| Vasopressor use, *n* (%) | 75 (50.3) | 97 (39.9) | 0.056 |
| Mechanical ventilation, *n* (%) | 110 (73.8) | 176 (72.4) | 0.853 |
| PaO_2_/FiO_2_ ratio, median (IQR) | 104 (77.0–154) | 107 (76.0–150) | 0.773 |
| Serum creatinine (mg/dl), median (IQR) | 1.61 (1.07–2.91) | 1.83 (1.08–3.30) | 0.211 |
| BUN (mg/dl), median (IQR) | 32.2 (22.0–53.7) | 35.0 (20.3–54.0) | 0.717 |
| Serum potassium (mEq/L), median (IQR) | 4.50 (4.00–5.30) | 4.50 (4.00–5.00) | 0.551 |
| Blood pH, median (IQR) | 7.34 (7.24–7.41) | 7.33 (7.25–7.40) | 0.904 |
| Blood bicarbonate (mmol/L), median (IQR) | 22.0 (19.5–25.1) | 23.0 (20.0–25.2) | 0.303 |
| D-dimer (ng/ml),^d^ median (IQR) | 3223 (1212–11008) | 3188 (1331–13974) | 0.888 |
| Hemoglobin (g/dl), median (IQR) | 12.7 (11.4–14.1) | 12.3 (10.9–13.8) | 0.045 |
| Leukocytes (cells/mm^3^), median (IQR) | 11,090 (7220–15,780) | 11,050 (7485–15,150) | 0.813 |
| Platelets (cells × 10^3^/mm^3^), median (IQR) | 205 (156–278) | 213 (160.5–287.5) | 0.43 |
| ALT (U/L),^e^ median (IQR) | 39.0 (26.5–63.0) | 36.0 (23.2–56.8) | 0.215 |
| CRP (mg/L),^f^ median (IQR) | 192 (95.9–300) | 209 (111–329) | 0.114 |
| **At KRT initiation** |  |  |  |
| Reason for KRT initiation |  |  |  |
| Hypervolemia,^g^ *n* (%) | 57 (38.8) | 74 (31.1) | 0.151 |
| Uremia,^g^ *n* (%) | 104 (70.7) | 166 (69.7) | 0.925 |
| Serum creatinine (mg/dl), median (IQR) | 4.36 (3.35–5.84) | 4.94 (3.69–6.56) | 0.02 |
| BUN (mg/dl),^h^ median (IQR) | 96.5 (69.5–122) | 93.9 (67.8–109) | 0.088 |
| **Outcomes** |  |  |  |
| Hospital stay (days) among survivors,^i^ median (IQR) | 45.0 (32.0–76.5) | 39.0 (22.0–63.0) | 0.176 |
| In-hospital death, *n* (%) | 130 (87.2) | 180 (74.1) | 0.003 |

KRT, kidney replacement therapy; CVVHD, continuous venovenous hemodialysis; IQR, interquartile range; BMI, body mass index; COPD, chronic obstructive pulmonary disease; eGFR, estimated glomerular filtration rate (by the 2021 Chronic Kidney Disease–Epidemiology Collaboration equation); N/A, not applicable; ICU, intensive care unit; SOFA, Sequential Organ Failure Assessment; PaO_2_, partial pressure of arterial oxygen; FiO_2_, fraction of inspired oxygen; BUN, blood urea nitrogen; ALT, alanine aminotransferase; CRP, C-reactive protein; CVVHD, continuous venovenous hemodialysis.

^a^Data available for only 372 patients (137 and 235 in the CVVHD and intermittent HD groups, respectively).

^b^Data available for only 156 patients (58 and 98 in the CVVHD and intermittent HD groups, respectively).

^c^Measured/determined within the first 24 h after ICU admission.

^d^Data available for only 363 patients (139 and 224 in the CVVHD and intermittent HD groups, respectively).

^e^Data available for only 365 patients (135 and 230 in the CVVHD and intermittent HD groups, respectively).

^f^Data available for only 368 patients (138 and 230 in the CVVHD and intermittent HD groups, respectively).

^g^Data available for only 385 patients (147 and 238 in the CVVHD and intermittent HD groups, respectively).

^h^Data available for only 385 patients (144 and 241 in the CVVHD and intermittent HD groups, respectively).

^i^There were 77 survivors (19 and 58 in the CVVHD and intermittent HD groups, respectively).
